# Consistency of sleep timing and duration are associated with more physical activity and favorable heart rate metrics in a naturalistic cohort

**DOI:** 10.64898/2026.06.09.26355325

**Authors:** Keyana Komilian, Inje Lee, Balaji Goparaju, Matt T. Bianchi

## Abstract

**Background:** Regularity of sleep patterns over time has increasingly gained traction as an important axis of sleep health. Since sleep habits are under some degree of behavioral control, understanding such patterns in naturalistic settings is particularly important. We quantified sleep variability and tested the hypothesis that regularity correlates with physical activity, resting heart rate (rHR), and heart rate variability (HRV).

**Methods:** We analyzed real-world digital health data from over 81,000 participants (over 18 million nights) who provided informed consent to participate in the Apple Heart and Movement Study and elected to contribute sleep, activity, and heart rate data to the study. Variability was quantified using the standard deviation (SD) computed from total sleep time (TST), sleep start time (S-start), end time (S-end), and midpoint time (MP), as well as the Sleep Regularity Index (SRI).

**Results:** The SD-based variability metrics correlated with one another (R values 0.74-0.92), and with the SRI metric (R values 0.62-0.64). More consistent sleep, by any metric, was associated with more activity and better rHR and HRV. The most consistent tertile for TST variability had higher median TST (6.9 vs 5.9 hours), more daily exercise (32.8 vs 20.4 minutes), lower rHR (62.4 vs 65.6 beats per minute), and higher HRV (40.6 vs 37.3), all p<1e-100. The findings were similar when variability was defined by S-start SD, S-end SD, MP SD, or SRI.

**Conclusion:** Sleep consistency metrics are highly correlated with each other, and consistency by any metric was associated with more activity, lower rHR, and higher HRV. While causality cannot be established, the results of this large, naturalistic observational cohort are consistent with the growing literature on the potential positive health associations of sleep consistency.

## 1. Introduction

Maintaining a regular sleep schedule is a cornerstone of sleep hygiene recommendations^1^. The importance of regularity was emphasized in a recent consensus statement from the National Sleep Foundation^2^, although the task force did not make specific recommendations for how to quantify the temporal characteristics of sleep over time. Consistency of timing also may enjoy the benefit of being an explainable and plausibly actionable way to maintain or improve sleep habits, although various external, work related, social, and behavioral factors may influence the extent to which someone can maintain a consistent schedule^3^, and circadian rhythm itself^4^, while clinical conditions such as insomnia may impact sleep habits and thus metrics of regularity.

Consistency or regularity of sleep has been studied in multiple research cohorts, where measures of increased variability is often associated with adverse health outcomes in cross sectional and prospective analyses. One review of longitudinal sleep tracking studies reported that standard deviation (SD) was the most common metric of variability reported^5^. Cohort studies typically use 7 or 14 nights of tracking, which is often based on traditional actigraphy. For example, in the UK Biobank cohort, sleep variability based on seven days of actigraphy data has been associated with mortality using the Sleep Regularity Index (SRI)^6,7^, with diabetes using the standard deviation (SD) of total sleep time (TST)^8^, and incident cardiovascular disease with TST SD^9^. Similarly, based on 7 nights of actigraphy in the MESA cohort, sleep variability has been associated with cardiovascular events ((TST SD and timing SD)^10,11^, metabolic markers (TST SD and timing SD)^12^, and adiposity (TST SD)^13^. Based on 14 nights of actigraphy, the TST SD was associated with CV risk in the CoLaus cohort^14^. Also using actigraphy, the Study of Osteoporotic Fractures showed variability in duration and in midpoint (both via SD) was associated with obesity^15^. Several recent reviews provide more comprehensive summaries of the extant literature^16,17^.

In the modern era of consumer sleep trackers, the opportunity arises to characterize sleep variability over longer time frames. We recently described the statistical impact of relatively short duration tracking when describing measures of centrality and dispersion in real and simulated sleep data^18^. In the current work, we sought to describe sleep consistency using several common metrics, and to explore potential associations with physical activity, resting heart rate (rHR) and heart rate variability (HRV) in a real-world longitudinal cohort of adult participants who consented to participate in the Apple Heart and Movement Study^19^. We hypothesized that the various measures of consistency would be correlated with each other, and that more consistent individuals by any variability metric would exhibit longer sleep duration, more activity, and improved cardiac metrics.

## 2. Methods

### 2.1 Apple Heart and Movement Study (AHMS) overview

AHMS was approved by the Advarra Central Institutional Review Board, and registered to ClinicalTrials.gov (ClinicalTrials.gov Identifier: NCT04198194). Informed consent was provided digitally via the Apple Research app. This study includes adults who consented to participate in AHMS^20^, and opted in to sharing HealthKit data streams including sleep tracking and other digital health data. This population consisted of adults residing in the United States who own an iPhone and an Apple Watch. AHMS collected a variety of data types including objective sleep and exercise data via HealthKit. Participants opted in to sharing data and could withdraw from the study at any time. The study ran from 2019 to 2025.

### 2.2 Study population

Although the study did not require or incentivize sleep tracking, many participants did track and opt in to sharing sleep data. The current analysis considered sleep data only when the data source was the first-party sleep application on Apple Watch, running iOS 16 and watchOS 9, or later. Sleep sessions were accepted using an operational rule for main sleep sessions (longest session if two or more sessions existed, overlapping with the participant sleep schedule).

The minimum data requirement for inclusion in the analysis of SD-based variability metrics was at least 7 nights of sleep data in the time window of November 2023 to March 2025, corresponding with the accelerometer based sleep stage classifier algorithm release and AHMS study end. The algorithm is a validated four-stage sleep classifier outputting awake, Core Sleep, Deep Sleep, REM sleep in 30 second granularity^21^). The mean number of nights per participant was 232. During this window, some participants joined the study, some withdrew, and the analysis includes all of those so long as they met the described sleep data availability inclusion criteria. For analysis of the SRI metric, at least one 30-day window with at least 15 nights of sleep data contributing to the SRI computation were required (mean of 27.6 nights). For the weekend versus weekday analysis, at least 10 weeks were required with both weekend nights (Friday and Saturday nights) and at least 4 weekday nights to be included. These criteria were pre-specified.

### 2.3 Data Types

The total sleep duration and time stamps of the start, end, and midpoint of the sleep period were obtained via HealthKit data for each night, using only Apple Watch first-party sleep tracking data. Activity data and heart data were likewise obtained from HealthKit, in the form of exercise minutes and energy expenditure (kCal), resting heart rate (rHR), and the SDNN form of heart rate variability.

The retrospective analysis was performed on data that was accessed on February 6, 2026, for this investigation. The authors did not have access to information that could identify individual participants during or after data collection.

### 2.4 Analysis of real-world health data

Statistical analyses and generation of plots were done in Python^22–24^. Correlations used the Spearman non-parametric method. Where p-values are incorporated, we used a 2-sided 0.05 threshold for significance. Sleep start was defined as the start time of the first epoch scored as sleep by the Apple Watch sleep stage classifier algorithm. The longest sleep period was used, unless a shorter session occurred within 60 minutes before the start time of the longest session, in which case the start time was taken as the preceding shorter session’s start time (this happened in <5% of nights). Sleep end was defined by the clock time of the last epoch of scored sleep in the primary (longest) sleep session. Sleep midpoint (MP) was defined as the middle of the clock-time window between start and stop of the primary sleep session. The SRI was computed using the formula from Fischer et al^25^. Nights with missing sleep data were ignored. For nights included in SRI processing, the surrounding time without sleep classifier output was classified as wake.

## 3. Results

### 3.1. Cohort description

**Table 1** provides a breakdown of the cohort, including demographics and wearable metrics for sleep, activity, and cardiac data types. The median number of nights of sleep per participant was 150, but the distribution was skewed toward higher values (such that the mean number was 232 nights). The distributions of TST, S-start, S-end, and MP values are given in **Supplemental Figure S1**.

**Table 1.** Participant Characteristics. Continuous variables are given as median (with 25th and 75th percentiles). BMI, body mass index; bpm, beats per minute; HRV, heart rate variability; M, male; F, female; ms, milliseconds; rHR, resting heart rate; WASO, wake after sleep onset. REM, rapid eye movement.

|  |  |
| --- | --- |
| N | 81,685 |
| Age | 46 (37, 58) |
| Sex (%) | M 63.8%, F 35.4% |
| BMI (kg/m²) | 27.2 (24.1, 31.3) |
| Active Energy (kCal) | 522 (369, 733) |
| Exercise (minutes) | 21 (7, 54) |
| rHR (bpm) | 63 (57, 70) |
| HRV (ms) | 37.9 (28.4, 50.6) |
| # nights per person | 150 (44, 377) |
| Total Sleep Time (minutes) | 404 (345, 453) |
| Core Sleep Time (minutes) | 270 (222, 314) |
| Deep Sleep Time (minutes) | 44 (31, 56) |
| REM Sleep Time (minutes) | 82 (59, 105) |
| WASO (minutes) | 17 (7, 40) |

### 3.2. Sleep variability defined by the standard deviation

To illustrate what kinds of patterns might be captured by various measures of sleep variability, **Figure 1** shows a series of simplified cartoon examples, and whether each metric would capture the forms of variability. For example, consider 3 nights of sleep, each with a single sleep period and no awake epochs. Holding the S-end constant, but letting the S-start vary, will not only read out in the S-start SD (and MP SD), but also in the TST SD because TST will vary in this cartoon example. Likewise, holding S-start constant, variability in the triad of nights would be captured by S-end SD, MP SD, and TST SD. In the example of altering both start and end, where the TST also changes, the MP is held constant, this will read out in variability of S-start, S-end, and TST. The final triad keeps start, end, MP, and TST constant, but varies the position of a wake bout. The SRI metric will capture variability by all sets of cartoon triads, including this last set, not captured by any of the SD metrics, although it is not typical clinically to describe variability purely from a wake after sleep onset (WASO) perspective.

**Figure 1.**
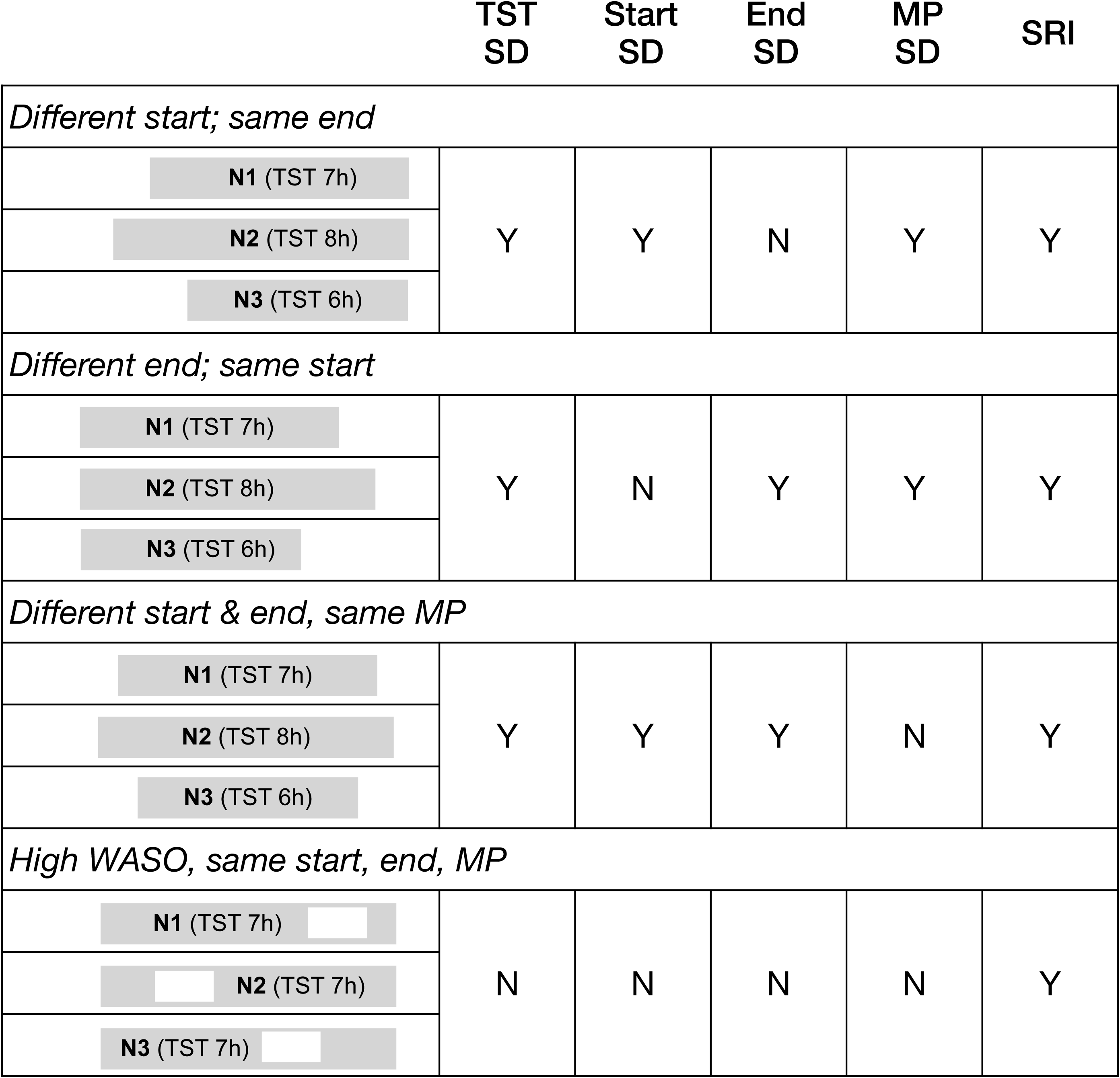
Cartoon illustrations regarding different metrics to capture variability. Three nights of sleep (gray horizontal bars, left column, N1-N3)) are shown for each example pattern, where the horizontal position corresponds to clock time and indicates relative position of the nights in time. The TST of each night is indicated in the corresponding gray bar. The italics text above each triad of nights describes the pattern shown. For each triad, the columns indicate whether each metric of sleep variability is predicted to capture the type of variability shown in the cartoon nights.

**Figure 2** shows the distribution of SD values for four metrics: Total Sleep Time (TST), Sleep Start (S-Start), Sleep End (S-End), and Sleep Midpoint (MP). The mean, median (p50), 25^th^ (p25) and 75^th^ (p75) percentile values are provided in the inset of each panel. The overall shapes are similar (longer tail skewing toward higher SD values), and the median values are similar except for the MP which was slightly smaller than the median SD value of the other three variability metrics.

**Figure 2.**
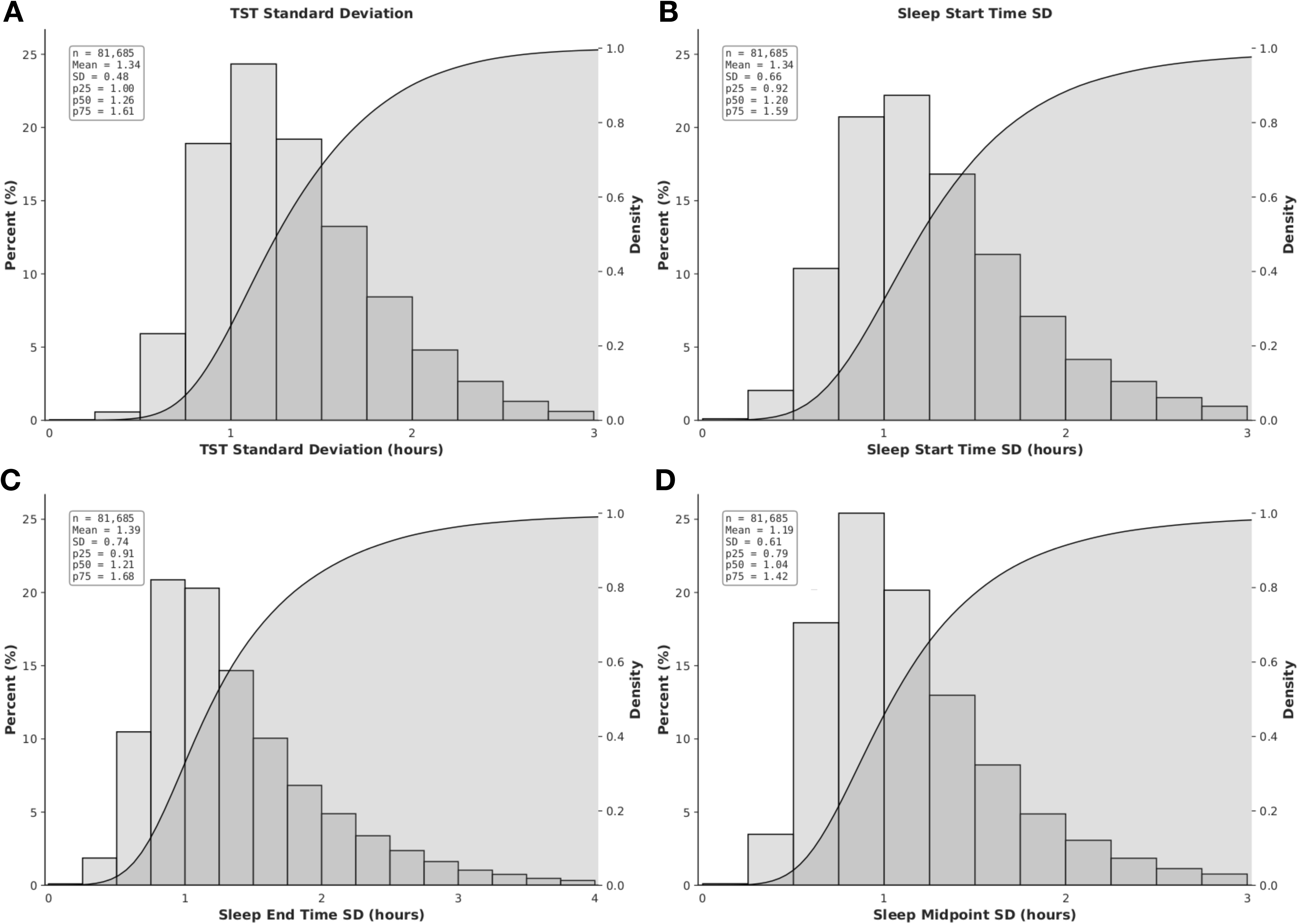
Distributions of four sleep variability metrics. The SD value was computed per participant for each of four variables: TST (A), S-start (B), S-end (C), and sleep MP (D). Each panel consists of a histogram view where the X axis values are bucketed, with the bar height values corresponding to the left Y axis, as well as a cumulative distribution function (S shaped solid line, with gray shading underneath), which corresponds to the right Y axis. The insets show the mean, SD, median (p50), 25th (p25) and 75th (p75) percentile values for each plot.

The distribution of SRI values in this cohort (**Figure 3A**) was computed using a different inclusion rule, given that it requires analysis of adjacent nights in time (see methods Section 2.2). The sample size remains large, since the time window was two years in which to capture qualifying 30 day windows. The median (IQR) of 77.4 (70.4, 82.4) was similar to that reported in the UKBB using actigraphy (median 81.0, IQR 73.8–86.3)^6^. The mean and median SRI values were similar if a more strict inclusion criteria requiring at least 15 adjacent nights was used (**Supplemental Figure S2**). We also computed SRI values using a one week window (see methods), which is commonly employed in epidemiological cohorts that include sleep tracking over time. **Figure 3B** shows the distribution of the difference (delta) between the two time windows, within-participant, to illustrate how different the SRI estimation can be when the sample window size is changed, similar to our recent work for standard statistical metrics of sleep over time^18^. While the mean and median were both within one point of each other, the 25^th^ and 75^th^ percentile span -4.30 to +4.65 points.

**Figure 3.**
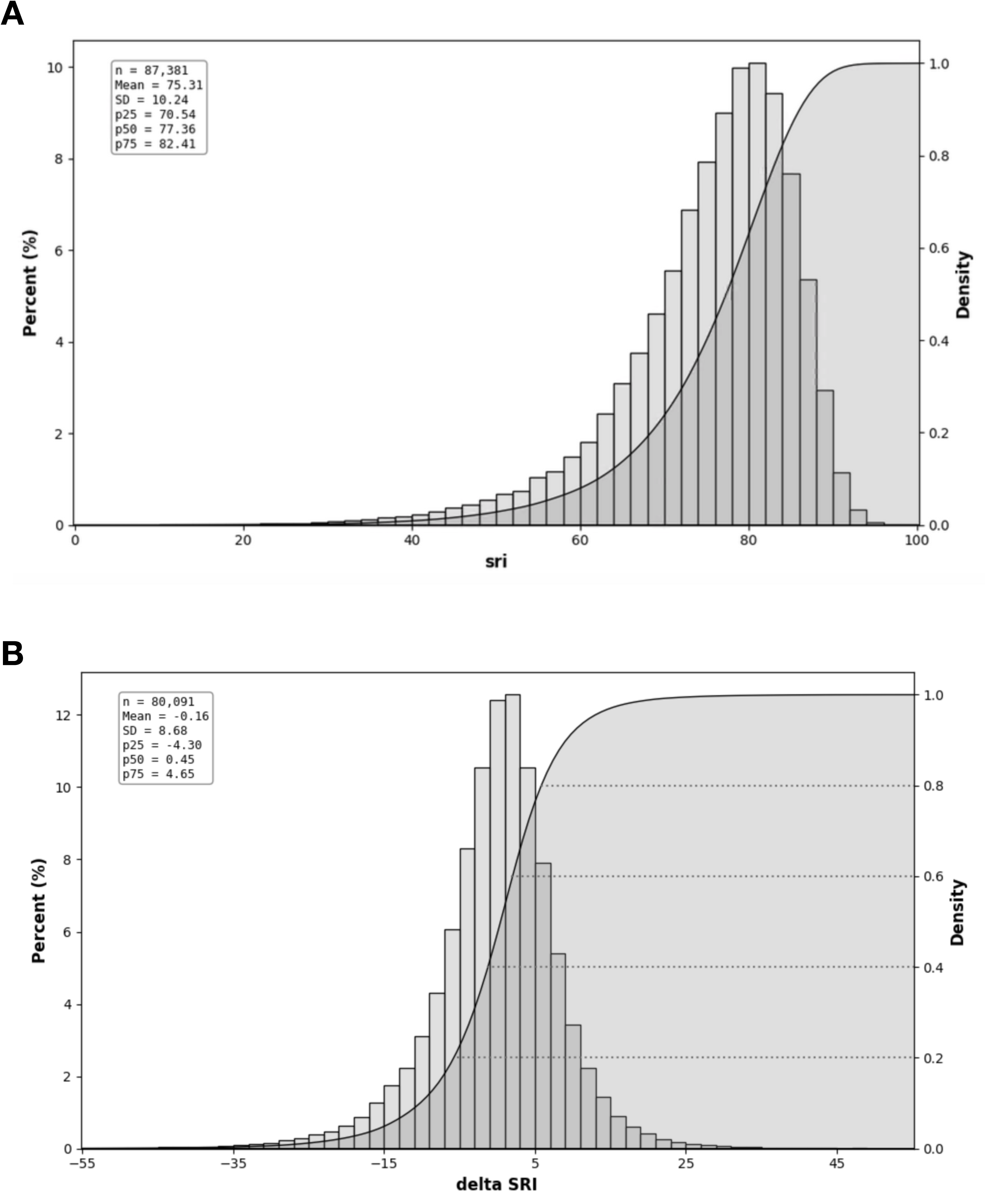
Distribution of the SRI metric in the cohort. The Sleep Regularity Index was computed for each individual based on a requirement to have at least 15 adjacent night pairs of sleep data in a 30 day window. In panel A, the histogram view buckets the X axis values, one computation per participant, with the bar height corresponding to the left Y axis, and is overlaid with a cumulative distribution function (S shaped solid line, with gray shading underneath it), which corresponds to the right Y axis. The insets show the mean, SD, median (p50), 25^th^ (p25) and 75th (p75) percentile values. In panel B, the delta between an SRI value computed from a 30 day time window is subtracted from the SRI value computed from a one week time window per participant (negative values means the week window had a lower SRI value).

Because SRI compares epoch by epoch from adjacent days, it is predicted to capture some similar aspects of variability compared to the SD of TST, S-start, S-end, and MP (as noted in the Supplemental data of Windred et al^6^), but as shown in **Figure 1**, it will also capture variability in how wake bouts occur over a night (as noted in the simulation study of Fischer et al^25^). To quantify the degree to which these metrics are capturing similar patterns of sleep in the real world, we computed a Spearman correlation matrix with all five values, as well as the mean TST and mean WASO values for context (**Figure 4**). The SD-based metrics were strongly positively correlated with one another, with R values from 0.74 to 0.92. The SRI metric was strongly negatively correlated with all SD-based metrics, with R values from -0.62 to -0.64.

**Figure 4.**
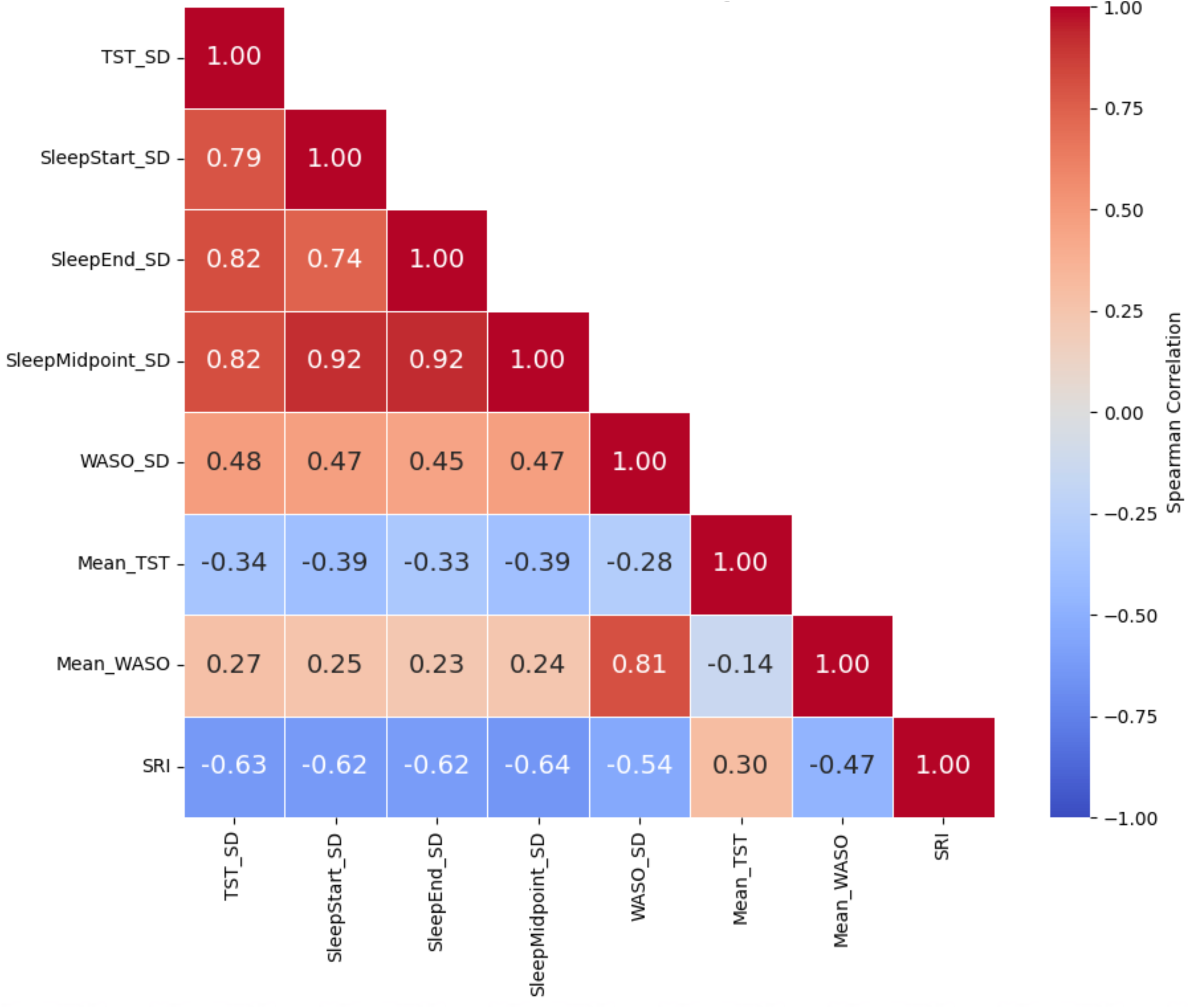
Correlation matrix of sleep matrix. The Spearman nonparametric correlation R value is shown in each cell of the correlation matrix, and also color coded for visual reference according to the scale at the right of the matrix. Each participant contributes one computed value for each metric, and the correlation is computed across the cohort. All R values are significant to p < 1e-100 . SD, standard deviation; SRI, sleep regularity index; TST, total sleep time; WASO, wake after sleep onset.

Of note, wake time during the night is not expected to align with clock time, and thus individuals spending more time awake in their sleep periods are predicted to have lower SRI values. The correlation matrix indeed shows this relationship, with R values of-0.54 with WASO SD, and -0.47 with mean WASO (**Figure 4**). We also observed that WASO is associated with higher values for the SD-based consistency metrics, more so for WASO SD (R = 0.45 to 0.48) than for mean WASO (R = 0.23 to 0.27). Finally, mean TST was associated with consistency, similarly for the SD based metrics (R = -0.33 to - 0.39) as for the SRI metric (R = 0.30).

### 3.3. Cross-sectional correlates of sleep variability

Having described a series of metrics designed to capture sleep variability over time, we next sought to investigate, cross-sectionally, if these metrics are associated with other digital health variables, including exercise minutes, kCal, rHR, and HRV. **Table 2** shows how these metrics are related to sleep consistency across the five sleep metrics used to characterize sleep consistency. The data are presented as the median and IQR values for the digital health metrics, according to whether the participant was placed in the top or bottom tertile for each sleep variability metric. Similar magnitude differences from most to least consistent tertiles were observed for each SD-based sleep variability metric: 11-12 minutes more exercise, 70-100 more kCal, 3 beats per minute lower rHR, 2-3 ms higher HRV, and about 1 hour more sleep duration. SRI tertiles showed similar magnitude delta values, although the HRV delta was higher at 5.2 milliseconds.

**Table 2.** Activity and heart metrics according to consistency tertiles. Participants were grouped according to their ranking of variability metrics by tertile. Exercise minutes, kCal, rHR, HRV, and TST were computed as mean values per participant, and those values are summarized as median and IQR values in each cell of columns two and three, representing the most consistent and least consistent tertile for each variability metric. The right column (Delta) is the subtraction of each row ’ s values in the least consistent from the most-consistent columns to allow comparison of how the digital health metrics associate with the different forms of sleep variability. All comparisons between most and least consistent are significant to p<1e-100, owing to the large sample size. rHR, resting heart rate; HRV, heart rate variability; kCal, kilocalories; ms, milliseconds; bpm, beats per minute; TST, total sleep time

|  | Most Consistent | Least Consistent | Delta |
| --- | --- | --- | --- |
| Exercise (minutes) |  |  |  |
| TST SD | 32.8 (18.5, 52.9) | 20.4 (11.2, 35.4) | 12.4 |
| S-Start SD | 31.9 (17.9, 51.9) | 20.6 (11.3, 36.0) | 11.3 |
| S-End SD | 31.6 (17.6, 51.7) | 20.4 (11.2, 35.3) | 11.2 |
| MP SD | 31.9 (17.8, 51.9) | 20.5 (11.2, 35.5) | 11.4 |
| SRI | 32.8 (18.7, 52.6) | 21.2 (11.7, 36.8) | 11.6 |
| kCal |  |  |  |
| TST SD | 571 (440, 730) | 469 (349, 617) | 102 |
| S-Start SD | 554 (427, 711) | 482 (359, 634) | 72 |
| S-End SD | 558 (432, 715) | 478 (355, 627) | 80 |
| MP SD | 556 (430, 712) | 482 (358, 633) | 74 |
| SRI | 567 (437, 725) | 492 (370, 640) | 75 |
| rHR (bpm) |  |  |  |
| TST SD | 62.3 (57.0, 67.7) | 65.7 (60.3, 71.6) | 3.4 |
| S-Start SD | 62.4 (57.2, 67.9) | 65.6 (60.1, 71.4) | 3.2 |
| S-End SD | 62.4 (57.2, 67.9) | 65.6 (60.2, 71.4) | 3.2 |
| MP SD | 62.3 (57.1, 67.8) | 65.6 (60.2, 71.4) | 3.3 |
| SRI | 62.3 (57.1, 67.7) | 65.4 (59.7, 71.3) | 3.1 |
| HRV (ms) |  |  |  |
| TST SD | 40.6 (31.9, 51.7) | 37.3 (28.8, 48.4) | 3.3 |
| S-Start SD | 40.0 (31.4, 51.0) | 37.7 (29.1, 49.2) | 2.3 |
| S-End SD | 40.5 (31.8, 51.5) | 37.8 (29.1, 49.2) | 2.7 |
| MP SD | 40.2 (31.5, 51.3) | 37.9 (29.1, 49.3) | 2.3 |
| SRI | 41.6 (32.8, 52.7) | 36.4 (28.1, 47.4) | 5.2 |
| TST (hours) |  |  |  |
| TST SD | 6.9 (6.4, 7.3) | 5.9 (5.1, 6.6) | 1.0 |
| S-Start SD | 6.9 (6.4, 7.3) | 5.9 (5.1, 6.6) | 1.0 |
| S-End SD | 6.8 (6.3, 7.3) | 5.9 (5.1, 6.6) | 0.9 |
| MP SD | 6.9 (6.4, 7.3) | 5.9 (5.0, 6.6) | 1.0 |
| SRI | 6.8 (6.4, 7.3) | 6.2 (5.5, 6.8) | 0.6 |

Another way to examine the association of consistency with digital health metrics is to label each participant as being in the highest (or lowest) tertile for 0, 1, 2, 3, or all 4 SD-based variability metrics. If we consider the most consistent group as being within the most-consistent tertile of all four SD-based metrics (N∼17k), and the least consistent group as being within the least-consistent tertile of all four SD-based metrics (N∼15k), we see somewhat larger separation of the digital health metrics (**Supplemental Figure S3**), compared to the univariate tertile method. For example, the mean exercise minutes was 34.2, versus 18.7 minutes, and the mean rHR was 61.8 bpm versus 66.1 bpm.

### 3.4. Weekend versus weekday sleep patterns

Given the possibility that weekend versus weekday behaviors may impact the computations used to describe sleep consistency, we analyzed each of the four SD-based sleep metrics accordingly. For this analysis we included only individuals who had at least 10 weeks in which sleep data was available on both weekend nights, and at least 4 weekday nights, to ensure adequate coverage for computing SD values (see Section 2.2)^18^. We observed that weekend nights were somewhat longer (mean of 6.8 vs 6.6 hours), and S-start times were shifted somewhat later (mean of 11:52pm vs 11:31pm) (**Figure 5**). The median value for TST SD was slightly lower on weekdays versus weekends (1.13 versus 1.19 hours for TST SD), and the median S-start SD was slightly lower as well (1.04 versus 1.16 hours) (**Supplemental Figure S4**).

**Figure 5.**
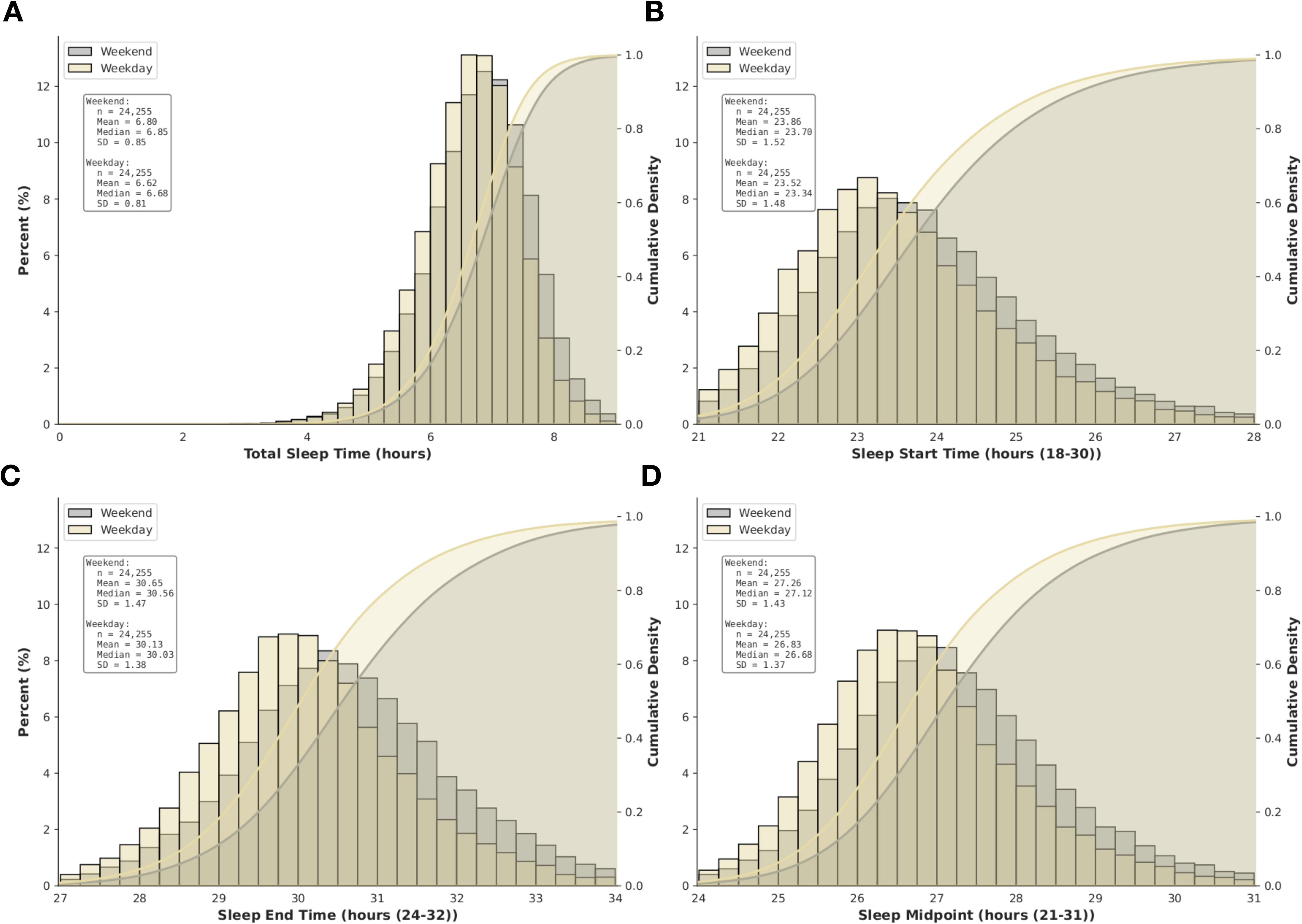
Distributions sleep duration and timing metrics. The mean value was computed per participant for each of four variables: TST (A), S-start (B), S-end (C), and sleep MP (D). Each panel consists of combined histogram and CDF, showing the data from weekdays versus weekends separately according to the color inset code. The insets show the associated summary metrics for each panel.

To further characterize the weekend versus weekday differences from a more individualized perspective, we computed for each participant the difference between the mean weekend and mean weekday values. The distribution of the “delta” between weekends and weekdays shows that most individuals are within +/- 30 minutes for TST (82.5%) and S-start (72.6%), while differences of greater than +/- 1 hour were less common (4.3% and 7.1%, respectively) (**Supplemental Figure S5**). It was more common to observe individuals with higher SD on the weekends than weekdays, perhaps not surprisingly (**Supplemental Figure S6**).

## 4. Discussion

This study describes sleep consistency metrics in a naturalistic, longitudinal cohort based on the Apple Watch sleep staging algorithm. The results demonstrated a high degree of correlation among four common methods of characterizing sleep variability: TST SD, S-start SD, S-end SD, and sleep MP SD. In addition, the more recently described SRI metric also shows high correlation with the four SD-based metrics, and a stronger correlation with WASO than the SD based metrics. Participants with more consistent sleep by any metric, compared to less consistent participants, showed higher exercise minutes and kCal, lower rHR, and higher HRV. Although weekend versus weekday differences were observed, they were not the driving factor these cross-sectional associations.

### 4.1 Measures of sleep variability are correlated with one another

Despite the acknowledged conceptual importance, there is no accepted standard method for quantifying the consistency or regularity of sleep over time^2^. It is not uncommon to see only one, or perhaps two, metrics computed from real world cohorts, most commonly the SD computed for the TST or the start time^5^. There are statistical, physiological, and behavioral reasons to suspect that measures of sleep consistency will in fact be correlated, leading to additional challenges regarding the choice of metrics for any given investigation. The statistical considerations regarding computation of a simple metric like the SD have been recently described, including the potential impact of weekend differences, especially when short duration tracking windows are used^18^. The multitude of behaviors^26–28^ intersecting with homeostatic and circadian processes lead to within and between individual differences that further challenge the invocation of any particular method to quantify variability. One approach to reconcile these challenges is a pragmatic one taken in the current study: whenever sleep consistency is assessed, different methods should be compared to avoid over-interpreting one measure in isolation. In this way, the combination of shared and independent aspects of variability could be recognized, and perhaps even separated depending on the study design.

### 4.2 Measures of sleep variability are correlated with activity and heart metrics

Cross sectional analysis revealed directional associations between more consistent sleep (by any metric) and more favorable activity metrics (higher exercise minutes and kCal values), lower rHR, and higher HRV. The digital health associations across the SD-based consistency metrics were highly similar, as was the case for the SRI metric, with the possible exception of HRV, which seemed to have a stronger association with SRI than the others. These associations are consistent with previous reports linking sleep with physical activity and heart metrics^29,30^. One potential explanation is that individuals who are able to maintain a consistent sleep schedule, which combines behavioral, external, and physiological factors, are also the types of individuals to maintain higher levels of physical activity, and those factors, either individually or in combination, drive the favorable rHR and HRV metrics. It is also possible that the consistent sleep habits lead to physiological benefits in autonomic function, in addition to the well established link between fitness levels and heart metrics. Despite these plausible hypotheses, causality is not established from the current analysis.

### 4.3 Considerations for epidemiology research

Aspects of sleep, from physiology and pathophysiology to patterns of timing and duration over time, have been associated with a variety of health outcomes. However, studies of temporal patterns may differ in methodology, including how sleep is measured (diary, actigraphy, wearable) and how variability is quantified (statistical methods of variation applied to the duration, timing, or across-day similarity of sleep). The current observations of sleep being longer and later on weekends versus weekdays have been reported using real world data from other consumer sleep trackers with different form factors and algorithms^31–34^. The wearable differences are smaller than that reported using mid-sleep time differences from survey data, which suggested >1 hour and >2 hour shifts on the weekends occur in 69% and 30% of respondents^35^.

While there are theoretical reasons to believe that different methods will be more or less capable of capturing certain patterns of variability (**Figure 1**), in real world data these methods are likely to correlate with one-another, as shown in the current study. This has implications for interpretation of study results in general, but also can impact statistical approaches, e.g., control variables assume a certain causal framework (e.g. treating confounding versus mediating factors differently).

The question has been posed whether SRI predicts outcomes better than TST, or if it predicts outcomes incrementally after controlling for TST. Our results suggest a distinct question which is in principle answerable from cohorts with outcomes data like the UKBB: to what extent does SRI (or any metric of sleep consistency) predict outcomes, compared to other metrics of sleep consistency. Although these metrics are clearly correlated with R values from 0.62 to 0.92, there is clearly unexplained variance among any given pair. Our analysis based on counting how many of the four SD metrics resulted in a low (or high) tertile assignment serves as one clue that there is some shared and some unique information among them.

Actigraphy algorithms typically exhibit high sensitivity for sleep but low specificity^36^. This operating point for binary classification predicts over-estimation of sleep in proportion to the amount of true wake in a night. This adds another angle to the interpretation of SRI, which is sensitive to the amount of wake between adjacent nights. Although there is no physiological limitation to when awakenings happen during a primary sleep session, we do observe that the mean as well as the SD of total wake duration are correlated with each other and with SD-based metrics of variability in TST, S-start, S-end, and S-midpoint (**Figure 4**).

In sum, the results would not favor, based on real world wearable data, a single “correct” or best way to capture sleep consistency. The NSF task force on this topic also did not recommend a particular method, noting the variety of methods that have been reported^2^. The current results highlight the overlapping and complex nature of real world sleep consistency metrics, suggesting that future research into sleep variability should attempt to quantify the incremental explanatory value when modeling health associations. For example, a hypothesis about SRI or TST SD should describe the unique or incremental explanatory value over other metrics of variability.

### 4.4 Limitations

Although the current report involves a large cohort under naturalistic conditions over time, there are limitations that could impact interpretation and extrapolation. For example, the extent to which the observations generalize to other populations, or to other consumer sleep tracking devices, remains uncertain. However, the correlation among SD-based metrics of sleep variability would not be expected to be a product of wearable algorithm performance, and the sleep metrics reported are similar to those obtained by actigraphy for example. The SRI metric may be more influenced by details of consumer tracker algorithms, which may differ in their capture of irregular sleep or napping behavior, to the extent that blocks of sleep outside a “primary” session are explicitly included in the SRI computation. Finally, although the cross-sectional associations of sleep consistency with favorable physical activity and heart metrics are physiologically plausible, it remains unknown if they are in fact causal.

## Data Availability

The study data and materials cannot be publicly shared due ethical and legal restrictions intended to protect participants privacy, honor the permissions granted by the participants in the informed consent and comply with Apple contractual obligations.

## 5. Acknowledgements

The authors acknowledge the important data contributions provided by all participants in the Apple Heart & Movement Study, without whom this research would not be possible. We also thank the following individuals for their thoughtful discussion, helpful guidance, and other valuable efforts in support of this work: Jeff Stein, Ian Shapiro, Nader Bagherzadeh, Dane Johnson, and Jonathan Varbel. The Apple Heart & Movement Study receives funding from Apple Inc. and the American Heart Association.

---

**Supplemental Figure S1.**
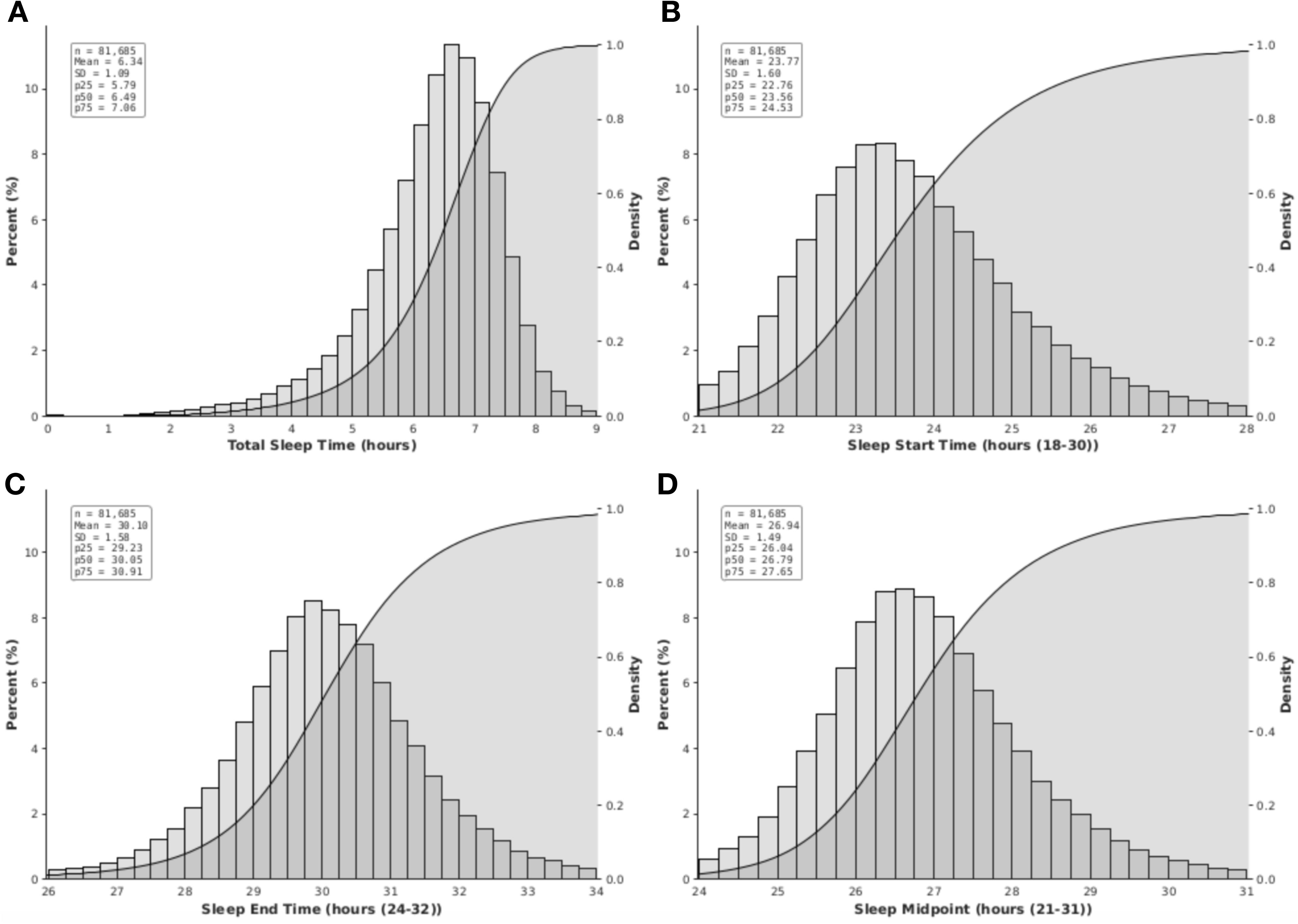
Distributions sleep duration and timing metrics. The mean value was computed per participant for each of four variables: TST (A), S-start (B), S-end (C), and sleep MP (D). Each panel consists of a histogram view where the X axis values are bucketed, with the bar height values corresponding to the left Y axis, as well as a cumulative distribution function (S shaped solid line, with gray shading underneath), which corresponds to the right Y axis. The insets show the mean, SD, median (p50), 25th (p25) and 75th (p75) percentile values for each plot.

**Supplemental Figure S2.**
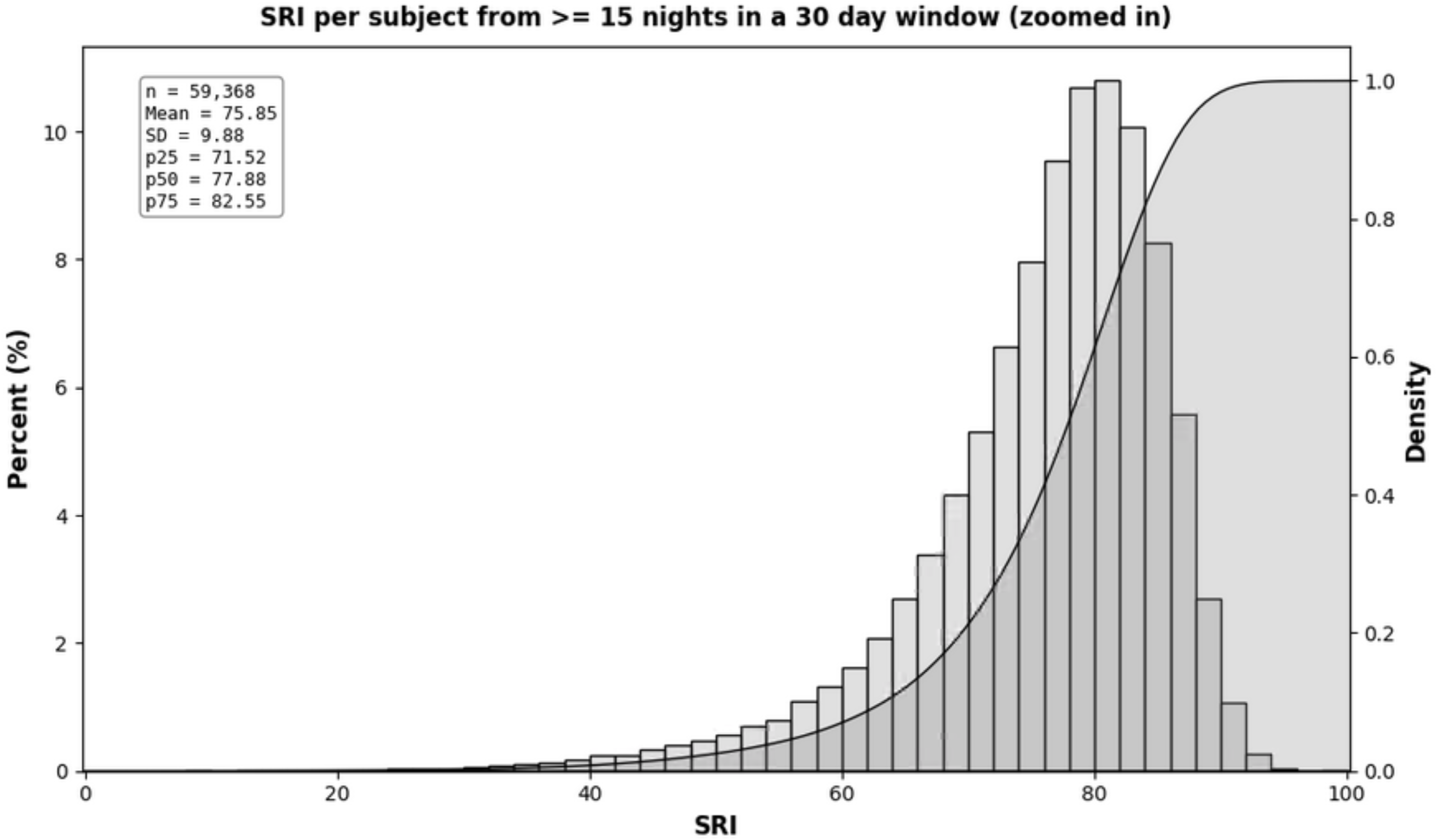
Distribution of the SRI metric in the cohort using a strict inclusion criteria for data availability. The Sleep Regularity Index was computed for each individual based on a requirement to have at least 15 adjacent nights (no gaps permitted) of sleep data in a 30 day window, a more strict criteria than that used for the distribution in Figure 3A. The frequency histogram (left Y axis) is shown overlaid with the cumulative density curve (right Y axis). The insets show the mean, SD, median (p50), 25^th^ (p25) and 75th (p75) percentile values.

**Supplemental Figure S3.**
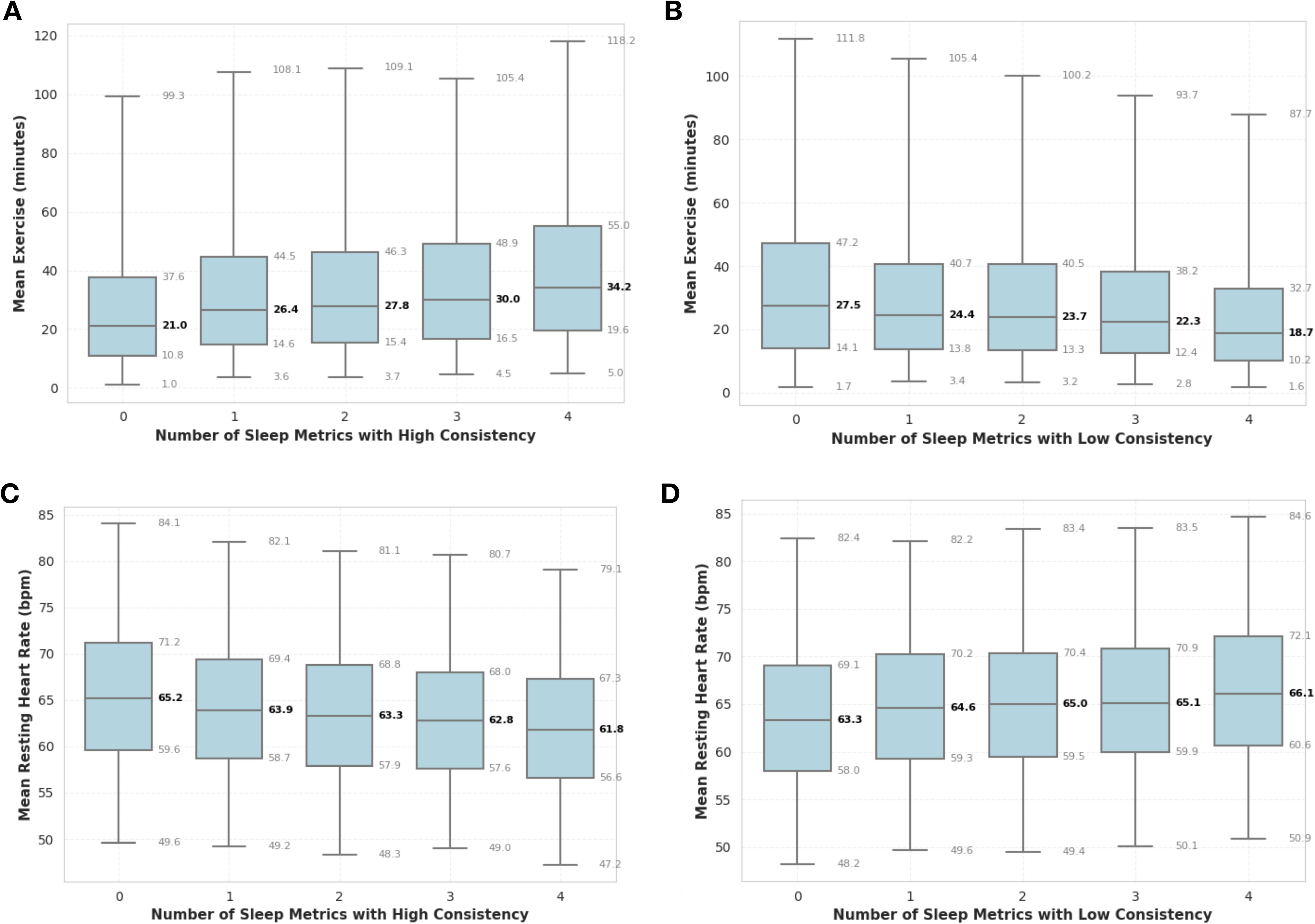
Activity and heart metrics associated with SD-based consistency metrics. The same data used in Table 2 were used in this analysis. The tertile assignment for each SD-based sleep consistency metric is used to assigned a count of number of metrics in the highest tertile (A and C) or lowest tertile (B and D). The mean exercise (A and B) and mean rHR (C and D) values are shown as box plots for each category, ranging from zero to four, corresponding to how many metrics the individual was in the top or bottom tertile. For example, in panel A, the zero category contains individuals who never were placed in the most consistent tertile for and SD-based sleep metric, while the 4 category contains individuals who were placed in the most consistent tertile in all 4 categories. The boxes in each panel correspond to the 25th, 50th, and 75th percentile, and the whiskers corresponds to the 5th and 95th percentile values (each number is shown adjacent to those markers).

**Supplemental Figure S4.**
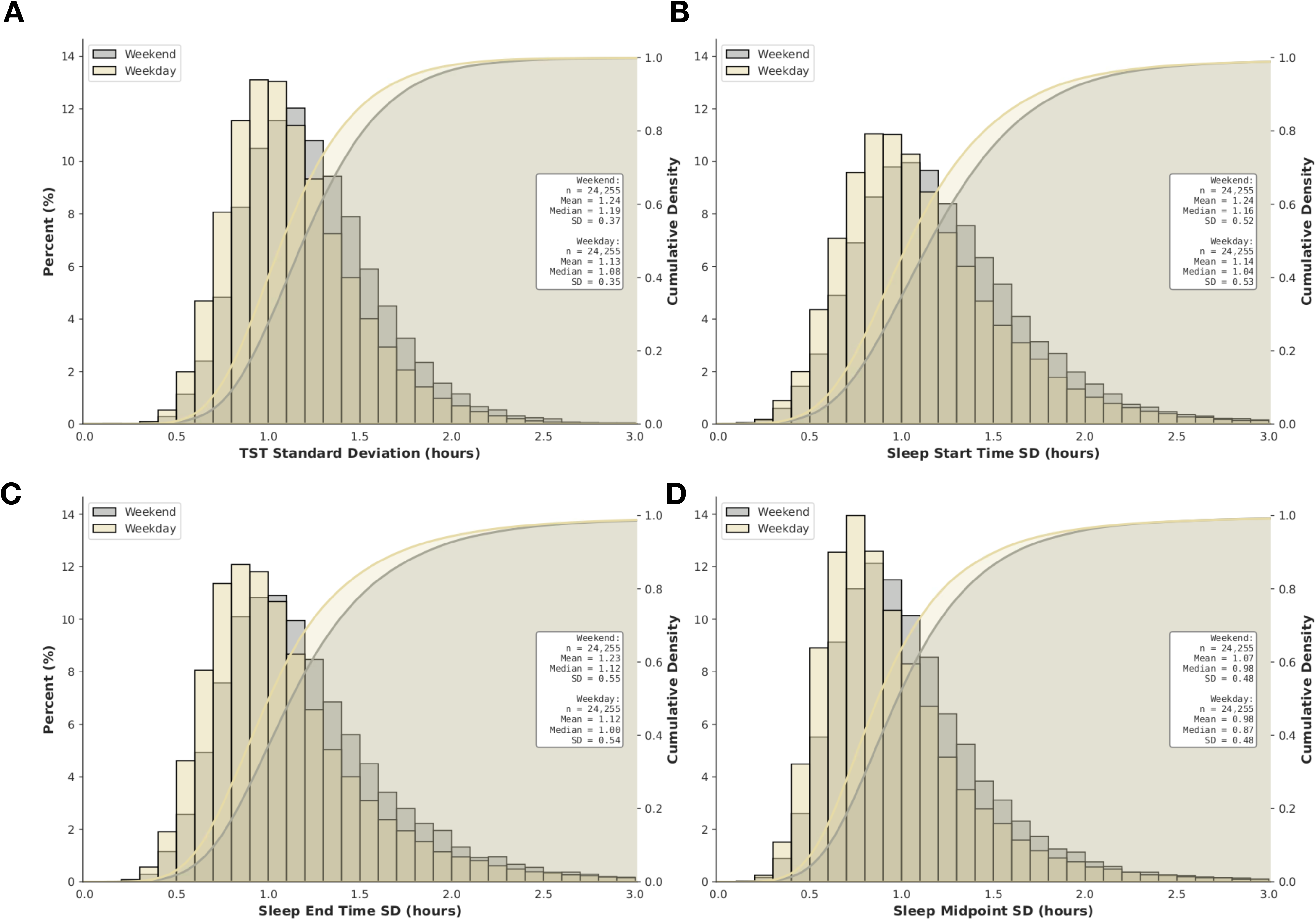
Distributions of SD-based sleep consistency metrics by weekday versus weekend. The mean value was computed per participant for each of four variables: TST (A), S-start (B), S-end (C), and sleep MP (D). Each panel consists of combined histogram and CDF, showing the data from weekdays versus weekends separately. The insets show the associated summary metrics for each panel.

**Supplemental Figure S5.**
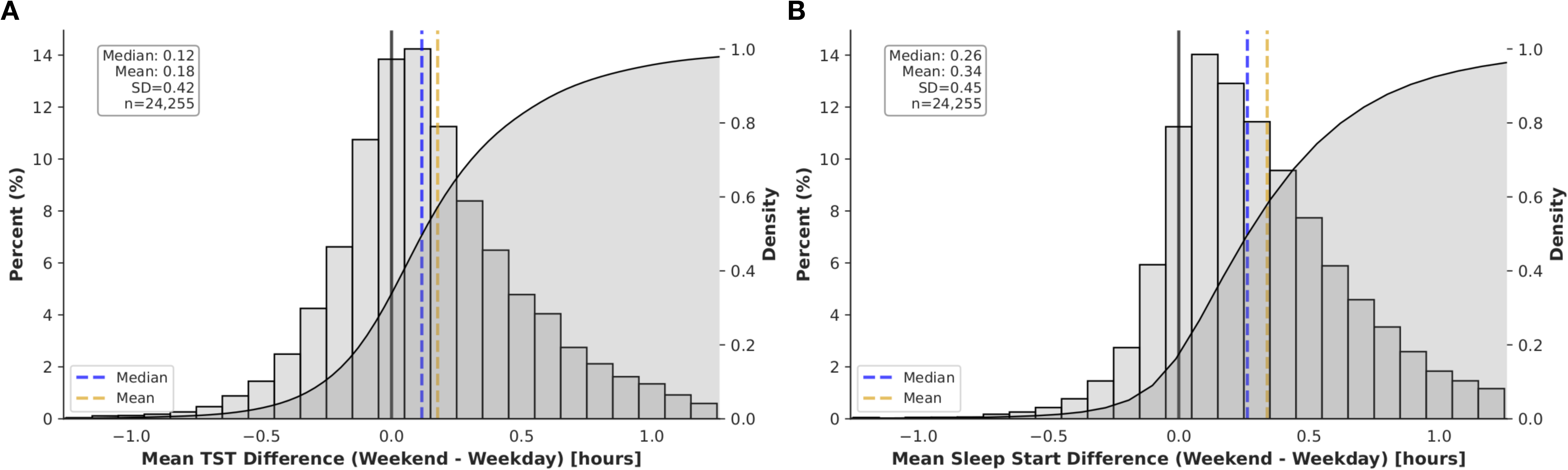
Weekend versus weekday differences, computed per-participant, in TST and S-start metrics. For each participant, the mean values for TST (A) and S-start (B) were computed, and the two values subtracted, such that positive values on the X axis in each panel mean higher weekend values compared to weekdays. The histogram view buckets the X axis values, with the bar height corresponding to the left Y axis, and is overlaid with a cumulative distribution function (S shaped solid line, with gray shading underneath it), which corresponds to the right Y axis. The insets show the mean, SD, median values.

**Supplemental Figure S6.**
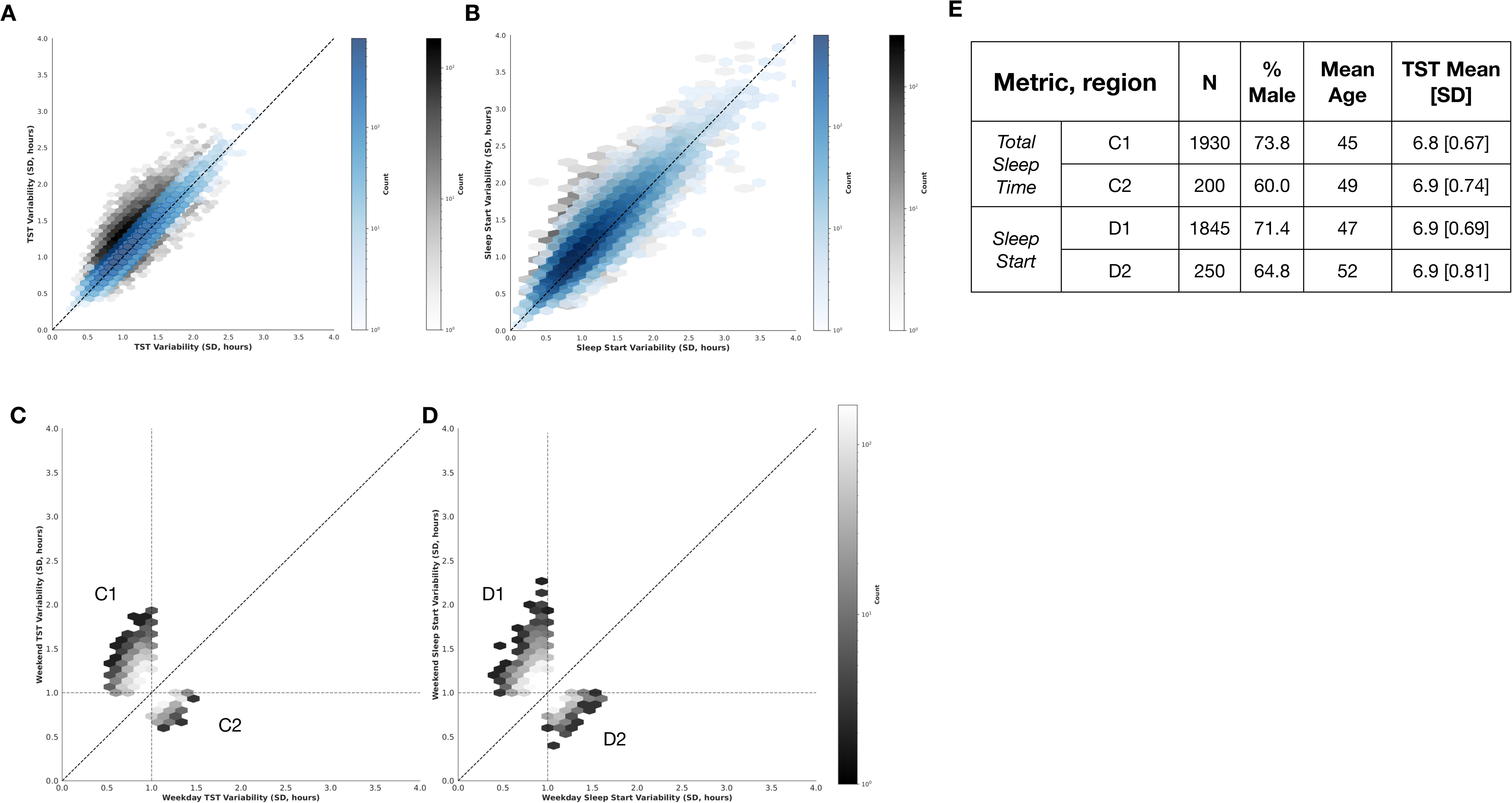
Weekend versus weekday variability, computed per-participant, in TST and S-start metrics. Panel A shows the SD values for TST for weekend nights (Y axis) and weekday nights (X axis). The diagonal dotted line is the identity line. Blue shading indicates individuals for whom the SD values are within 15 minutes of the identity line, and the sample size is in the blue legend; areas outside this “tunnel” are in gray scale and correspond to the grayscale legend. Panel B follows the same format, showing variability in sleep start by weekend vs weekday. Panels C and D correspond to a subset of panels A and B, showing only “regions” demarcated by a 1 hour SD value on each axis, excluding the blue shaded tunnel region. Panel E shows the sample size, age, sex, and mean TST value for those in the “discordant” regions indicated.

